# Knowledge and experiences of community health workers in the delivery of physical rehabilitation services in Zambia- a qualitative interview study of trained community health workers

**DOI:** 10.1101/2023.06.12.23291153

**Authors:** Miriam Mapulanga, Thembelihle Dlungwane

## Abstract

**Aim:** To explore the knowledge and experiences of community health workers in the delivery of physical rehabilitation services in Zambia.

**Design:** An exploratory qualitative study

**Setting:** Lusaka district, Zambia

**Participants:** Ten community health workers from the Zambia Enhanced Community-Based Rehabilitation Program

**Results:** Nine major themes emerged from the data. Four of these themes (patient management, treatment programs, issuing of appropriate assistive devices through a multidisciplinary approach, and levels of care) were highlighted as the required knowledge base of community health care workers (CHWs), whereas five themes (patient assessment and treatment, home program and family education, assistive devices issuing, a well-functioning referral system, unrealistic caregiver expectations, and resource constraints) were revealed as the essential experiences of CHWs who delivered the required physical rehabilitation services. The CHWs under study experienced unrealistic expectations from caregivers, such as the high expectation that physical rehabilitation would heal a child. These expectations were complicated by a lack of understanding among the caregivers of the role and capabilities of CHWs. Moreover, constraints such as limited resources and a lack of permanent employment were perceived as major challenges.

**Conclusion:** Trained community health workers have adequate knowledge and experience in delivering physical rehabilitation services in areas where more specialised and qualified physical rehabilitation workers do not exist. In light of the current shortage of qualified physical rehabilitation workers in Zambia, more investments are required, particularly in the training of community health care workers, to increase physical rehabilitation service coverage.

**KEY MESSAGES:** *What is already known regarding this topic:* Community health workers have been involved in the delivery of physical rehabilitation services. They are therefore key in improving the quality of life of the general population and that of disabled people through the various roles that are task-shifted to them by trained and qualified physical rehabilitation workers. However, their knowledge and experiences in delivering physical rehabilitation services remained relatively unknown until the current study on which this paper is based.

*What this study adds to the pool of knowledge:* The findings affirm that, with appropriate training in physical rehabilitation, community health workers can effectively deliver physical rehabilitation services and issue appropriate assistive devices at community level in Zambia.

*How this study might affect research, practice, and policy:* The study proposes that appropriately trained community health care workers be utilised to effectively deliver physical rehabilitation services at community level where such services by appropriately qualified physical rehabilitation workers are lacking.

## BACKGROUND

The World Health Organisation (WHO) describes community health workers (CHWs) as “lay care health workers selected from the community where they operate and are answerable to the very communities” that they serve [1]. In many low-income countries (LICs) like Zambia, CHWs have become the backbone of the community health system as they are an integral part of primary health care (PHC) and are essential for achieving universal health coverage (UHC) [2, 3], particularly in rural and poor communities. Community health programs have been extensively used to address the chronic shortage of human resources for health (HRH) through task shifting, which is an adaptive system that leads to improved access to health care and improved health outcomes [4, 5]. More specifically, CHWs have been involved in the delivery of physical rehabilitation services and have thus been involved in improving the quality of life of disabled people through task shifting various roles to them by trained physical rehabilitation workers like physiotherapists and occupational therapists [6, 7, 8]. In settings where such programs have been implemented, CHWs deliver physical rehabilitation services to the community as rehabilitation health workers or rehabilitation assistants [9, 10].

It is estimated that there are fewer than ten skilled rehabilitation workers per one million people in LICs, and this results in population’s limited access to rehabilitation services. The shortage of HRH therefore has an adverse effect on communities in LICs amid a high burden of chronic and communicable diseases that can lead to physical impairments [11, 12, 13]. In countries where CHWs have been used to deliver physical rehabilitation services, they have adopted various roles such as case managers, providers of basic assistive devices and issuing assistive devices [14, 15, 16]. Physical rehabilitation task shifting has also resulted in the training regimen of CHWs ranging from two-day sessions to a two-year training program that covers various physical rehabilitation components like health promotion, exercise prescription, and palliative care [14, 17].

Zambia’s National Health Care is responsible for physical rehabilitation services that are provided at three levels [18]. With a population of 19 million, Zambia is plagued by a HRH crisis hence the need for an increased ratio of HRH to general population [19, 20]. While Zambia’s healthcare system is decentralized, the majority of appropriately qualified physical rehabilitation workers are found in urban areas and in settings that are accessible by railway. The majority of these HRH are located in three main areas, namely Lusaka, the Copper Belt, and Southern provinces [18, 21]. Due to a dire shortage of physical rehabilitation workers, primary health care level is particularly affected, hence non-governmental organizations (NGO) supplement physical rehabilitation services at community level by engaging a few physical rehabilitation workers and CHWs. Therefore, these services provided are mainly aligned with the relevant NGO’s objectives, such as treating children with cerebral palsy and ensuring that the disabled are provided with orthotics or prosthetics [22].

Task shifting physical rehabilitation services to CHWs at the primary health care level improves accessibility, especially for the disabled and the vulnerable [23]. However, while the physical rehabilitation services that are delivered by CHWs need to be safe, efficient, effective, equitable, and sustainable, there is no standardised approach to the training of the CHWs who are tasked with physical rehabilitation services [24]. Therefore, understanding CHWs’ knowledge and experiences regarding physical rehabilitation service delivery is essential. The purpose of this paper was to investigate the knowledge and experiences of CHWs trained in physical rehabilitation and thereby deliver physical rehabilitation services in the selected study area in Zambia.

## METHODS

### Study design and setting

An exploratory qualitative case study approach was used to investigate the knowledge and experiences of CHWs trained in physical rehabilitation and thereby deliver physical rehabilitation services in Lusaka. The study was conducted in Kanyama, an informal settlement, three kilometres **west of Lusaka city centre.** The study follows a constructivist paradigm and results are presented adhering to the SRQR reporting guideline for qualitative research [25].

### Patient and public involvement

No patients involved.

### Sampling procedure and study participants

Purposive sampling was employed to select CHWs who were trained in physical rehabilitation services under the Zambia Enhanced Community Based Rehabilitation Programme (ZECREP). The programme aims at improving the quality of life of children with disabilities in Zambia by 2030. While the project is run by Cheshire Homes Society of Zambia with the support of the Liliane Foundation of Holland, one of its local partners is the Archie Hinchcliffe Disability Intervention (AHDI) which operates in Lusaka district. Therefore, the study recruited all the ten CHWs who were trained by instructors from the AHDI for ten days in November 2020. Their training focused on physical rehabilitation service delivery under the ZECREP project.

### Data collection

Data were collected by means of a face-to-face in-depth interview with each of the participants in April 2022. The interview schedule was developed based on the objectives and the ultimate aim of the study, hence it posed open-ended questions that facilitated prompting. The objectives of the study, that focused on physical rehabilitation services were to determine CHWs’ knowledge regarding physical rehabilitation service delivery and to explore what experiences the CHWs had of physical rehabilitation service delivery. The topics were covered that could elicit data on CHWs’ knowledge and experiences of physical rehabilitation service delivery with specific focus on patient assessment, treatment plans, home visits, family education, use of assistive devices, and referrals. All the interviews were conducted in English by the principal investigator at the health facility where the CHWs were supervised. Each interview was audio recorded and transcribed verbatim. The completeness and quality of the transcripts were verified using observational notes that had been taken during the interviews. The principal investigator listened to the recordings and read the transcriptions repeatedly and then made a summary. Each interview lasted 45 to 60 minutes. In this study, data saturation was achieved when no new themes emerged.

### Data analysis

The transcribed data were managed and processed using Nvivo version 12. The participants were de-identified using codes, with first participant being identified as P1. The data were analysed using thematic content analysis to describe the interview content and to interpret the findings by drawing on the emerging themes [26]. Analysis commenced with the principal investigator reading and re-reading the interview transcriptions several times for familiarization. The aim and objectives of the study were considered throughout this process. This repeated reading process was followed by creating codes and categories that were grouped and classified as subthemes. The process continued until all the subthemes had been regrouped under themes. Eventually, the emerging themes and subthemes, along with their supporting quotes, were critically analysed and evaluated.

### Trustworthiness

Trustworthiness and scientific rigour were maintained throughout the study. Credibility was ensured by engaging in debriefing sessions with the supervisor. A peer qualitative researcher was also engaged to scrutinise the audio recordings and the transcribed data. By engaging this independent researcher in a peer reviewing process, the transcribed data from the interviews could be checked against the audio recordings to enhance the accuracy and quality of the data, and thus dependability was ensured. Transferability was assured by the presentation of detailed descriptions of the background of the study and the methods that were utilised. The study setting, sample selection and composition, and the data collection and analysis processes are therefore clearly and unambiguously described to establish the context and viability of the study and to allow for comparison. Confirmability was ensured during the study by exposing the transcribed data to validation and confirmation processes to ensure the accuracy of the transcripts used for analysis. Moreover, the study supervisor provided input throughout and reviewed the themes to ensure the reliability of the data analysis and interpretation processes.

### Researcher characteristics and self-reflection

MM is a physical rehabilitation provider and a public health practitioner as well as a health worker trainer. TD is a physical rehabilitation provider and a public health practitioner as well as an educational researcher. All the study participants were unknown to the researchers prior to the study.

## RESULTS

Ten interviews were conducted with purposively selected CHWs. All the participants had been trained in physical rehabilitation service delivery under ZECREP. The CHWs’ ages ranged from 34 to 56 years. The highest education level attained by the participants was Grade 12, while the lowest qualification was Grade 7. The CHWs’ years of experience in general community health were between 5 to 15 years, and all the participants had been delivering physical rehabilitation services for a minimum of 16 months before the interviews (Table 1).

**Table 1:** Profile of the study participants.

| **Code number** | **Sex** | **Age** | **Total Years' Experience in Community Health Service Delivery** | **Highest education level attained** |
| --- | --- | --- | --- | --- |
| 1 | Female | 31-35 | 6 | Grade 12 |
| 2 | Female | 36-40 | 5 | Grade 7 |
| 3 | Male | 45-50 | 10 | Grade 9 |
| 4 | Female | 35-40 | 7 | Grade 9 |
| 5 | Female | 41-45 | 12 | Grade 7 |
| 6 | Female | 46-50 | 9 | Grade 8 |
| 7 | Female | 35-40 | 5 | Grade 10 |
| 8 | Female | 51-55 | 10 | Grade 7 |
| 9 | Female | 41-45 | 6 | Grade 9 |
| 10 | Male | 56-60 | 15 | Grade 7 |

### Themes and Subthemes

Nine major themes emerged from the data. Four of the themes (patient management, treatment programs, issuing of appropriate assistive devices, and multidisciplinary approaches and levels of care) emerged as the knowledge of the participating CHWs regarding the delivery of physical rehabilitation services, whereas six themes (patient assessment and treatment, home program and family education, assistive device issuing, a well-functioning referral system, unrealistic caregiver expectations, and resource constraints) emerged as the experiences of the CHWs who delivered physical rehabilitation services. (Table 2).

**Table 2:** Themes and subthemes.

| **Objectives of the Study** | **Emerging themes** | **Subthemes** |
| --- | --- | --- |
| To determine the knowledge of CHWs regarding the physical rehabilitation services they rendered | Patient management | History taking<br>Problem assessment<br>Treatment focus on the problem |
|  | Treatment programs | Schedule<br>Specific day per child<br>Educating the entire family |
|  | Issuing of appropriate assistive devices | Use of home-made devices |
|  | Multidisciplinary approach and levels of care | Referring patients to other levels of care<br>Interacting with other rehabilitation health professionals |
| To explore the experiences of the CHWs regarding the physical rehabilitation services they rendered | Patient assessment and treatment | Appropriate diagnoses<br>Providing appropriate treatment<br>Anxieties related to the correct diagnosis |
|  | Home program and family education | Home activities<br>Involving everyone in caring for the child |
|  | Issuing assistive devices | Making devices for use<br>Importing sophisticated devices |
|  | A well-functioning referral system | Good turnaround times<br>Continuity of care |
|  | Unrealistic expectations among caregivers | High expectations that physical rehabilitation will heal the child.<br>Lack of understanding of the role of CHWs |
|  | Resource constraints | Limited resources<br>Lack of permanent employment |

### Addressing Objective 1: Knowledge of CHWs regarding physical rehabilitation

#### Patient management

The CHWs assessed children by obtaining information from their mothers related to the history of the affliction and the severity of the condition. One participant stated:

> *We get information from the mothers about the history of the condition, and the child’s abilities and inabilities. Then we do our assessment to see where the problem is. (P7)*

This comment suggests that the CHWs had sufficient knowledge to check the severity of the child’s condition and to determine appropriate treatment goals. Another participant added the following:

> *We also look at the child’s abilities, [such as] if the child can eat or sit… We see what he is not able to do, and that’s how we assess. And that is when we know where to start. (P8)*

Another participant stated:

> *When you are assessing a child, you want to know how the child got the disability and find out the problems the child is facing. Sometimes you find that the child did not cry at birth, then you’ll know that’s what caused the disability. (P6)*

At the end of the assessment, the CHWs and the child’s caregiver/s would agree on the treatment plan for the child. The CHWs selected suitable days for visitation and progress assessment. A participant discussed this process as follows:

> *Only after assessment, do we then know what is wrong with the child [and] we make plans for how we are going help the child. I show them activities to do too, and then I tell them the days I will be visiting them to check on what they are doing and to do my activities. We agree that they will be doing activities in the morning or afternoon depending on when the mother is free. (P1)*

Although the prescribed treatments seemed to focus on the children’s disabilities, their treatment plans also offered guidance on educational teaching activities, handling techniques, and nutrition. One participant stated the following:

> *There are things that these children can do and things they are unable to do. So, our focus is on what the child is unable to do. If the child is unable to speak properly, we teach communication…For those children who are unable to stand and sit on their own, we teach the mother lifting techniques. We also teach caregivers how to prepare nutritious food because some children with cerebral palsy have difficulties eating, resulting in malnutrition. (P3)*

#### Treatment programs

The therapy the children received was home-based as determined by the programme that was followed, therefore weekly home visits by the CHWs were deemed essential to ensure progress. These home visits were dependent on the CHWs’ respective schedules and a specific day and time were allocated for each home visit. One respondent said:

> *We only see our patients at home because here at the clinic there is no program for the disabled. So we only make home visits based on a schedule. (P10)*

Another participant corroborated this process by stating:

> *I make rounds visiting the patients based on a schedule. Each child has a specific day allocated to them when I go to assess how they are faring. (P8)*

The objective of home visits was to involve the caregiver and family in the treatment program of the child. One participant added the following:

> *We manage our patients at their homes with the parents involved. (P2)*

The CHWs also educated the household members on how to care for the disabled child. A participant explained this process as follows:

> *After the assessment, we ask if the family members know about the child’s condition and, if they don’t, we educate them so that the entire household knows how to care for the disabled child in case the primary caregiver is not available. (P1)*

#### Issuing appropriate assistive devices

The CHWs created assistive devices to help the disabled children they treated and to enhance the rehabilitation progress. They admitted that these devices were simple and homemade and that they used local materials to make the activities the children engaged in sustainable. A participant offered the following information in this regard:

> *We make balls for children to play with. But if the child is very young, we make toys that can hang above them [mobiles] so that they try to reach out to them for stimulation. (P6)*

To encourage and stimulate mobility, homemade bars were also made and used, as one participant explained:

> *If a person is unable to walk on their own, we make parallel bars [that are use] at their homes. (P3)*

#### A multidisciplinary approach and levels of care

It was found that the CHWs did not work in isolation as they made referrals to physiotherapists who would then assist those children in dire need of help. They revealed that referrals were made depending on the severity of the needs of the child involved. A participant stated:

> *If we cannot manage the condition, we have to refer that child to a hospital. (P1)*

Referrals were reportedly made for various reasons, but most particularly in cases that the CHWs could not manage, as the following participant explained:

> *If a child needs advanced physiotherapy, we refer him or her to a physiotherapist. Those who need social support or cash transfers are referred to social welfare. We work well with these people and they know all the programs. (P3)*

### Addressing Objective 2: The experiences of CHWs with physical rehabilitation service delivery

#### Patient assessment and treatment

According to the CHWs, it was important to make assessments before arriving at a correct diagnosis as only then would the way be paved for appropriate intervention. Various emotions were admittedly experienced during the assessment process, as one respondent explained:

> *After I have assessed a child I tell the parents what caused the disability of that child. Some parents will know what caused the disability, whether it was asphyxia or prolonged labour. Assessment is like a door that leads you somewhere, but you have to pass through so that you know where you are going. (P7)*

Another participant said:

> *When you assess very well, you’ll be able to help them to manage or you will refer them if necessary. The process can be distressing, but it requires that you take your time so that you arrive at the right diagnosis. (P1)*

While the CHWs were generally hopeful of the treatments they prescribed, they also experienced stress and despair following some diagnoses. For example, a participant shared the following:

> *Sometimes I am filled with hope during the assessment, but sometimes am filled with desperation depending on the condition and the diagnosis I have arrived at. I hope that when I have the diagnosis, then we can try to make the life of the child better. I become quite desperate if the diagnosis reveals that the outcome is not very hopeful for the parents. (P3)*

#### Home programmes and educating the family

The CHWs generally had a positive attitude regarding the home and family education programmes they initiated because they argued that educating all the other family members encouraged them to get involved in caring for the afflicted child, and therefore the child would be treated in the safety of the home. One participant stated:

> *With the education we are doing, if the mother is unable to do it, that child will be cared for by other family members. So we let all family members know what needs to be done to care for the child and at which particular time it needs to be done. (P8)*

Another participant said:

> *Family education is a good approach because if the whole family is knowledgeable, the burden will not be on one person alone. In case the primary caregiver is not around, the patient can still be cared for by other household members. (P9)*

#### Issuing assistive devices

It was generally agreed that assistive devices greatly helped both the patient and the caregivers as their use ensured some hands-off time for the caregivers and some independence for the patient. The need for an assistive device was established during the initial assessment session or during the physical rehabilitation process. When the need for a device had been established, the CHW would make simple toys using local materials like old used clothes, plastic, or cardboard of different colours. One participant said:

> *We make toys, those simple toys, like plastic balls and dolls using plastic and old clothes. (P3)*

The CHWs felt they were not skilled in constructing complicated assistive devices and therefore they had to depend on external suppliers in cases where sophisticated and technologically advanced devices were required. One participant described her experiences as follows:

> *We make simple devices like children’s seats using cardboard and cassava flour on our own, but with regards the complicated ones like walkers, we have to be supplied. (P4)*

#### A well-functioning referral system

It was admitted that some children needed to be referred for more advanced treatment. In this regard, the CHWs narrated positive encounters as they felt that their referrals were needed and effective because the children were attended to promptly despite some congestion at the facilities where they worked. A participant offered the following comment:

> *Children are usually attended to in time when we refer them even if there is congestion. (P6)*

Another participant stated:

> *When we refer a child, the referral is fruitful. If you refer a child for further assistance, the child will be attended to. This gives me satisfaction because we know that they will see that we’re improving the child’s condition for the future. (P1)*

#### Unrealistic expectations among caregivers

It was clear that some caregivers had unrealistic expectations of the CHWs. For instance, some parents expected a child to start walking shortly after treatment had started, and this impacted negatively on the mental state of the CHWs as they feared that they would never meet that expectation. One participant expressed her dissatisfaction with such unrealistic expectations as follows:

> *Some parents want to know how many days it will take for the child to walk before they commit themselves to the program. When we go and see the child, some parents expect quick improvement. So it makes me fearful of visiting those homes because of these [unrealistic] expectations. (P3)*

Moreover, the participants revealed that some parents expected more than physical rehabilitation support from the CHWs. A participant described this as follows:

> *When we visit some people, they think they will receive material [financial] help from us, but we explain to them that we do not offer material help. This affects how they receive us in their homes. (P1)*

#### Resource constraints

The CHWs highlighted that they were challenged by resource constraints such as inadequate schools and inappropriate infrastructure for children with disabilities. A participant discussed these constraints as follows:

> *Not having special needs schools is disheartening because the child’s progress is then hindered by a lack of education. Accessing education for a disabled child is a challenge. Many disabled children cannot access facilities because they were built poorly with no consideration for the disabled. There are no ramps to assist the mobility of a wheelchair, so it is a matter of lifting [and carrying] the person for him to enter the church or school L and it’s the same for toilets. (P10)*

The CHWs also lamented the lack of permanent employment opportunities which they felt posed a challenge in terms of the sustainability of their services to the community as well as their financial income to support their families. A participant explained this challenge by stating the following:

> *We are not [permanently] employed, so it is a challenge to balance this voluntary work and our various activities that help us to support our families. (P1)*

The lack of permanent employment clearly had an impact on their ability to take care of their families as well as their motivation to continue their support for disabled children. A participant stated:

> *We lack financial motivation. Yes, we are volunteers, but we also have to provide for our families at home. (P6)*

## DISCUSSION

This show that with training CHWs’ had gained adequate knowledge to diagnose the severity of disabled children’s condition and to prescribe some treatment procedures to support them, but this knowledge was clearly not enough in more severe cases as such children would be referred to more appropriately trained professionals. They had relatively similar experiences regarding the physical rehabilitation services they provided, such as the creation of simple assistive devices and educating the entire family to support the disabled child in the household. The literature affirms that the performance and skills of CHWs that render physical rehabilitation services are essential for the benefit of the patient, the caregiver, and themselves [27]. Moreover, research has shown that trained CHWs who engage in rehabilitation programmes can retain knowledge and improve their service delivery significantly [28]. As a sustainable solution to the shortage of physical rehabilitation workers, the knowledge and experiences of CHWs are important in the community health care field [29].

The participants highlighted that they were knowledgeable of patient management, planning treatment programs, issuing simple assistive devices, and in diagnosing and referring patients who require more sophisticated treatment regimens. The CHWs had adequate knowledge to take a patient’s history. They did this by obtaining information from the child’s caregiver/s and they conducted problem assessment by examining the child’s abilities and inabilities. In history taking, obtaining information from caregivers is a crucial part of the assessment process as it contributes to making the correct decision about the child patient’s needs [30]. Additionally, the CHWs could design and focus on treatment regimens for the children in their care and they could devise treatment plans that incorporated activities that would enhance the life of their patients and improve the involvement of the family and caregivers. Generally, the CHWs operated on guidelines used by qualified providers who assess children subjectively before an objective physical examination to ascertain the child’s problems and determine the most appropriate rehabilitation procedure [31].

The CHWs’ mode of delivery was home-based, which is an effective rehabilitation approach in improving the functioning of disabled children [32], especially with cerebral palsy children. While the CHWs scheduled a specific day of the week per child for treatment amidst other healthcare duties, literature shows that increasing treatment intensity results in better outcomes for children as treatment frequency should be established not on the principles of rehabilitation and child development, but rather on clinical reasoning and scheduling [33, 34]. Meanwhile, the CHWs offered health education to the family to equip them all with knowledge and insight about the cerebral palsy child’s condition [35]. This approach was for families to enhance their knowledge and skills, and fostered household education in the care of disabled children.

The fact that the CHWs made and issued home-made assistive devices to the disabled children in their care is in line with Articles 4, 20, and 26 of the Convention on the Rights of Persons with Disabilities, which requires countries to make devices available to the disabled at low cost [36]. Weak health systems in low-income countries such as Zambia pose a dire challenge regarding assistive device availability due to resource constraints and a lack of trained personnel which, in turn, leads to low domestic production and availability of assistive devices. The onus in such countries thus falls on CHWs to provide low-cost aids to assist the population, particularly children, who need support [37, 38].

The CHWs were knowledgeable of the primary health care system and how to refer patients to other health professionals who were members of the multidisciplinary health care team. While the CHWs’ mode of operation was home-based, they were supervised by a trained physiotherapist who was based at the zonal health centre. This centre was reportedly primarily responsible for health promotion and disease prevention and some limited curative services, while complicated cases were referred to the Level 1 hospital [18]. This means that, whenever cases required further management, the CHWs made referrals and did not need to work in isolation. The literature affirms that a functioning process of referrals ensures the continuation of case management which is a vital process for quality health service delivery [39].

However, the CHWs experienced anxiety when they were required to make a physical examination and clinical diagnoses using information provided by the children’s caregivers to arrive at an appropriate diagnosis. As members of the communities they served, the CHWs strove to provide appropriate treatment regimens that would be culturally acceptable and beneficial to their young patients. The World Health Organisation endorses this approach in a systematic review which states that CHWs should provide culturally acceptable services while improving service access for communities [40]. The clinical diagnoses the CHWs made were thus dependent on their competence which had been instilled during the training they had received. This suggests that any training CHWs receive should be of an appropriate standard to guide them to assess a patient and arrive at a correct diagnosis [41].

The data concerning physical rehabilitation service delivery by CHWs revealed that their experiences were diverse. For instance, some of these experiences related to patient care while others included patient/caregiver expectations and a lack of resources. A common feature was that all the CHWs conducted home-based care programs and family education, which means that their services were embedded in families at community level and that family education was core to their practice. This was a heartening finding as the involvement of family members in health interventions, particularly where disabled children are concerned, is associated with a positive influence on family health practices, and thus knowledge at household level of the management of a disabled child is recommended [42].

In their efforts to assist the children in need of special care, the CHWs made simple tools that could be used as assistive devices by the children. However, they admitted that they had to access more sophisticated devices via relevant health authorities. Their dedication thus meant that the use of simple assistive devices by their young patients was assured, while external support for more complicated ones was accessible. The goal of the WHO Community-Based Rehabilitation Guidelines on Assistive Devices is that disabled people should have access to suitable devices to help them participate in everyday life [43]. The role of health systems is therefore to determine which assistive devices are needed by which disabled persons, and to facilitate access to such devices. Furthermore, health systems are mandated to ensure the maintenance, repair, and replacement of such devices when necessary [43]. However, as the access to assistive devices in LICs like Zambia are limited, homemade assistive devices have been a solution to help with children’s progress in physical rehabilitation [37].

The CHWs also had positive experiences of the referral process as they reported prompt attention to the children who needed more advanced care. This is commendable as it speaks to the fact at primary health care, child health is a health priority in Zambia [18]. Apart from the fact that preferential health treatment is an enabler of a good referral system, the proper organisation of health services enables service providers to work as a team, and this team spirit facilitates effective referrals from one point to the other [44].

On a less positive note, the CHWs were exposed to unrealistic expectations from the caregivers of the children they served. For instance, there was the high and unrealistic expectation that physical rehabilitation would heal the child, and quickly. In health service delivery, such unrealistic expectations among caregivers may be attributed to anxiety and insufficient information [45], and this underscores the need for family health education programmes. Additionally, due to the community’s lack of understanding of the role of the CHWs, they expected to receive financial benefits from them. This is unfortunate and may be attributed to the fact that some NGO-driven programs actually provide material benefits for families.

The CHWs were also constrained by a lack of resources which negatively impacted the physical rehabilitation services they were able to deliver. For instance, the lack of schools for special needs children and inappropriate infrastructure for disabled children in existing schools limited the outcomes of their work as their clients could not progress optimally. Zambia, like many other developing countries, is challenged by a lack of special education facilities and infrastructure for the disabled as a result of poor political will, cultural influences, and limited resources that are all compounded by poverty [46]. Moreover, the fact that these CHWs were volunteers who were not remunerated meant that they lacked the necessary resources to support their own families. The literature also notes that a lack of permanent employment opportunities for CHWs is a barrier in various CHW programmes, particularly as most programs in sub-Saharan Africa are donor funded and mostly for vertical disease programs [47]. Integrating CHWs, like those who participated in the study, into the national health care system like any other salaried health worker will therefore guarantee permanent employment and the sustainability of their contribution to health care services, especially for disabled children.

### Limitations

This qualitative study was conducted in Lusaka, Zambia. Therefore, transferability of the study findings to other settings is limited. The focus of the study was on CHWs trained in physical rehabilitation services and did not include those that were not trained in delivering physical rehabilitation services.

## CONCLUSION

The knowledge of the CHWs regarding the physical rehabilitation of disabled children was enhanced through training as they were able to understand basic patient management, plan treatment programs, create and issue appropriate assistive devices, and work in multidisciplinary teams. However, their experiences regarding patient care, patient expectations, and the availability of resources were somewhat diverse, while they all agreed that their experiences of patient referrals were positive. The knowledge base and experiences that are highlighted in this paper may guide the development of a more suitable training model for CHWs that will enhance their knowledge, skills, and development and that will ensure their professional integration into the health care system in Zambia. In consideration of the shortage of committed and properly trained HRH in low-income countries, it seems vital that CHWS are adequately trained in physical rehabilitation service delivery on a national scale to curb existing shortages.

## Data Availability

All data produced in the present work are contained in the manuscript

## ABBREVIATIONS

WHO: World Health Organisation
CHWs: Community Health Workers
UHC: Universal Health Coverage
HRH: Human Resources for Health
ZECREP: Zambia Enhanced Community-Based Rehabilitation Program
OVC: Orphans and Vulnerable children
SSA: Sub-Saharan Africa
LICs: Low Income Countries
AHDI: Archie Hinchcliffe Disability Intervention

## Affiliation

University of KwaZulu-Natal, School of Nursing and Public Health, Department of Public Health Medicine, 2^nd^ Floor George Campbell Building, Durban 4001, Republic of South Africa

## Acknowledgements

We would like to acknowledge Ms Getrude Kapulisa, the Director of Archie Hinchcliffe Disability Intervention, and Mr Michael Chilufya Mutafya, the CBR focal point, for their support during the data collection process.

## Contributors

MM conceptualised and wrote the manuscript. TD supervised and contributed to the writing of the manuscript.

## Patient consent for publication

Not applicable.

## Ethical approval

This study was cleared by the University of KwaZulu-Natal Biomedical Research Ethics Committee (BREC 00000569/2019), and permission to conduct the study was obtained from the Zambia National Health Research Authority.

Miriam Mapulanga ORCID ID - http://orcid.org/0000-0001-5519-6635

## Participants’ consent for publication

Participants had the right to opt out of the study without any repercussions. Confidentiality was maintained throughout the study by using codes instead of real names for study participants and data were not made available to any other partner or project for further analysis. A consent form was signed by the participants before the interview in which they agreed that the data they provided could be used anonymously for research and publication purposes.

## Data availability statement

No data available

